# Discovering early imaging biomarkers of osteoradionecrosis in oropharyngeal cancer by characterization of temporal changes in computed tomography mandibular radiomic features

**DOI:** 10.1101/2020.10.09.20208827

**Authors:** Souptik Barua, Hesham Elhalawani, Stefania Volpe, Karine Al Feghali, Pei Yang, Sweet Ping Ng, Baher Elgohari, Robin C. Granberry, Dennis S. Mackin, G. Brandon Gunn, Katherine A. Hutcheson, Mark S. Chambers, Laurence E. Court, Abdallah Mohamed, Clifton D. Fuller, Stephen Y. Lai, Arvind Rao

**Affiliations:** Department of Electrical and Computer Engineering, Rice University, Houston, TX, USA; Department of Computational Medicine and Bioinformatics, University of Michigan, Ann Arbor, MI, USA; Department of Radiation Oncology, The University of Texas MD Anderson Cancer Center, Houston, TX, USA; Department of Radiotherapy, European Institute of Oncology, Milan, Italy; Department of Radiation Physics, The University of Texas MD Anderson Cancer Center, Houston, TX, USA; Department of Head and Neck Surgery, The University of Texas MD Anderson Cancer Center, Houston, TX, USA; Department of Oncologic Dentistry and Prosthodontics, The University of Texas MD Anderson Cancer Center, Houston, TX, USA; Department of Radiation Oncology, University of Michigan, Ann Arbor, MI, USA; Department of Radiation Oncology, Peter MacCallum Cancer Centre, Melbourne, Victoria, Australia

## Abstract

Osteoradionecrosis (ORN) is a major side-effect of radiation therapy in oropharyngeal cancer (OPC) patients. In this study, we demonstrate that early prediction of ORN is possible by analyzing the temporal evolution of mandibular subvolumes receiving radiation. For our analysis, we use computed tomography (CT) scans from 21 OPC patients treated with Intensity Modulated Radiation Therapy (IMRT) with subsequent radiographically-proven ≥ grade II ORN, at three different time points: pre-IMRT, 2-months, and 6-months post-IMRT. For each patient, radiomic features were extracted from a mandibular subvolume that developed ORN and a control subvolume that received the same dose but did not develop ORN. We used a Multivariate Functional Principal Component Analysis (MFPCA) approach to characterize the temporal trajectories of these features. The proposed MFPCA model performs the best at classifying ORN vs Control subvolumes with an area under curve (AUC) = 0.74 (95% confidence interval (C.I.): 0.61-0.90), significantly outperforming existing approaches such as a pre-IMRT features model or a delta model based on changes at intermediate time points, i.e. at 2- and 6-month follow-up. This suggests that temporal trajectories of radiomics features derived from sequential pre- and post-RT CT scans can provide markers that are correlates of RT-induced mandibular injury, and consequently aid in earlier management of ORN.

## Introduction

Radiotherapy (RT) is a highly utilized modality in the treatment of head and neck (H&N) cancers with well-established local control and survival benefits [1]. Advances in radiation delivery techniques from 2-dimensional (2D) and 3-dimensional (3D) techniques to intensity modulated radiotherapy (IMRT) with ability to manipulate the beam path to spare normal tissues has significantly improved cure rates and toxicity profile [2]. Despite that, osteoradionecrosis is a late complication from radiation to the mandibular bone with a serious impact on quality of life for a growing population of younger surviving head and neck cancer patients [3]. The incidence of ORN varied between different modalities ranging from 2-40% in the conventional era to 0-6% in the IMRT era. Different risk factors were identified to play a role in the development of ORN following radiotherapy treatments [4, 5]. Osteoradionecrosis has great impact on the patients’ life quality if not detected and managed properly [6, 7]. Diagnosis of ORN mainly relies on clinical and radiological tools such as computed tomography (CT) and magnetic resonance imaging (MRI) with their limited capacity for early detection [8].

Fortunately, the recent advances in biomedical imaging were coupled with the rise of radiomics in terms of extracting quantifiable imaging features, possibly of high information yield and subsequent computation of these features kinetics (e.g. delta-radiomics) derived from sequential images [9]. Paired with machine learning techniques, we hypothesize that radiomic feature kinetics can characterize and distinguish mandibular bone subvolumes at higher risk of developing future ORN. These “temporal virtual digital biopsies” might have the potential to empower earlier intervention and hence improve patients’ quality of life.

Consequently, the aims of this study are to:

1. Determine bone radiomic features derived from contrast-enhanced CT (CECT) images that are significantly different between ORN and non-ORN mandibular subvolumes.
2. Develop a composite radiomics-based signature integrating inputs from multiple pre- and post-treatment time points; with potential predictive utility of ORN incidence in high-risk mandibular subvolumes.
3. Hypothesis generation for future prospective studies.

## Materials and Methods

### Study Population

Following an approval from an institutional review board (IRB) at our institution, data for biopsy-proven OPC patients treated between 2002 and 2013 who underwent radiation therapy as a single or multimodality definitive therapy were considered for the current investigation (n=83). This investigation and relevant methodology were performed in compliance with the Health Insurance Portability and Accountability Act (HIPAA) as a retrospective study where need for informed consent was waived [10]. Electronic medical records were scanned for documented diagnosis of mandibular ORN following IMRT in the absence of any prior head and neck re-irradiation along the same lines as a previous ORN study by our team [11]. The aspects of our institutional IMRT approach for oropharyngeal cancer patients were previously reported in detail [12]. All patients received pre-radiotherapy Dental Oncology service clearance, and, if indicated, prophylactic dental extraction and fluoride trays, were prescribed as per standard Head and Neck Service operating procedure [13]. Inclusion and exclusion criteria for patients’ selection are illustrated in **Fig 1**.

**Fig 1.**
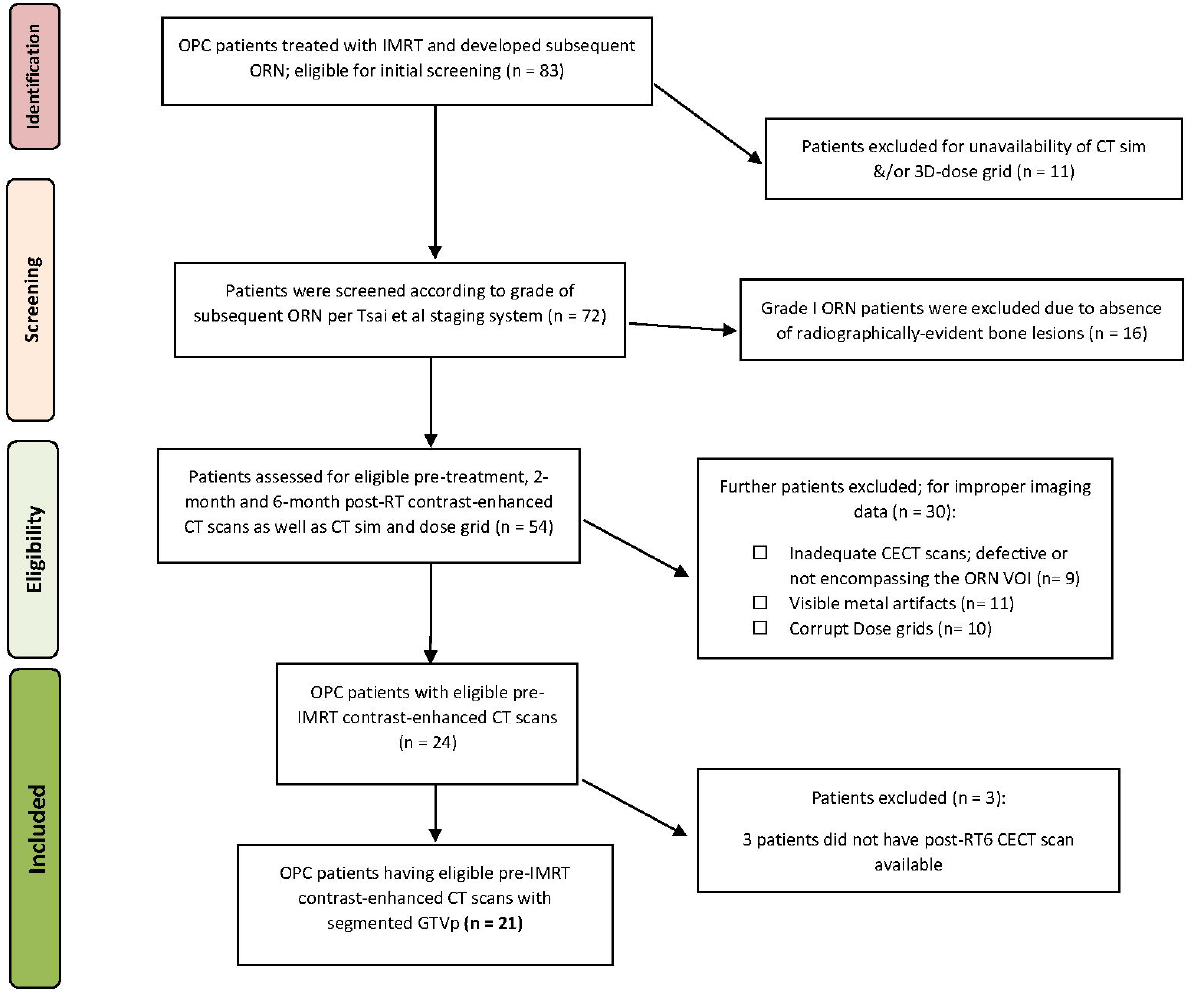
Patient selection. Flowchart of selection process of patients for this study

### ORN staging

The severity of ORN was graded I through IV as follows: grade I, i.e. minimal bone exposure requiring conservative management; grade II: minor debridement required; grade III: hyperbaric oxygen therapy (HBOT) received; grade IV: major surgery mandated. This staging system is very comprehensively given its emphasis on response to treatment as a standard to categorize ORN[13]. Patients who subsequently suffered from radiographically &/or pathologically-proven grade II or worse ORN were included in this study.

### CT Acquisition Protocol & eligibility criteria

According to our institutional protocol, CECT images were obtained as a prerequisite for pre-treatment diagnostic work-up. Subsequent post-IMRT CECT scans for response evaluation and further surveillance were routinely performed at 2-month and 6-month time points and then at regular preset intervals thereafter. Our study revolved about extracting quantitative imaging biomarkers from CECT at pre-IMRT (i.e. baseline), 2-month (post-RT2), and 6-month (post-RT6) post-IMRT, as well as time instance corresponding to development of ORN. To that end, CECT scans with available non-reconstructed axial cuts at the aforementioned 4 time points were retrieved. CT slices with evident ORN lesions that were obscured or otherwise affected by visible metal artefacts were not contoured and were not included in analysis.

All CT scans were attained with a multi-detector row CT scanner. Scan parameters are as follows: slice thickness reconstruction (STR) ranges between 1 and 3 mm, with median STR of 1 mm, X-ray tube current of 99-584 mA (median: 220 mA) at 120-140 kVp. All images acquired at our institution were composed of 512 x 512 pixels and were acquired following a 90 second delay after intravenous contrast administration. One-hundred and twenty milliliters of contrast were injected at a rate of 3 ml/sec. To standardize the image voxel sizes for use in texture feature calculations, all the CT scans were resampled, via a trilinear interpolation voxel resampling filter [14].

### Image Segmentation & Registration

We specifically selected CECT scans demonstrating the earliest radiographically evident ORN characteristic lesion(s) as reported by radiologists and further confirmed by physical examination by physicians in Head & Neck Surgery as well as in Dental Oncology [ORN CECT].

The original delivered DICOM-RT clinical treatment plans were restored from Pinnacle treatment planning system (Pinnacle, Phillips Medical Systems, Andover, MA) into commercially available image registration software (VelocityAI™ 3.0.1). Diagnostic CECT scans at baseline, post-RT2, post-RT6, and ORN were also imported. Radiographically evident bony lesions were delineated manually by a radiation oncologist (HE) to constitute the ORN volumes of interest (VOIs). Physical exam and other available imaging modalities such as dental-dedicated panoramic X-rays were utilized to guide the segmentation of VOIs.

Planning CT was co-registered with ORN CECT using deformable image registration algorithm of VelocityAI™ 3.0.1. The 3D reconstructed dose grid of RT plan was then overlaid to the ORN CECT. A neighboring radiographically intact mandibular subvolume within the same isodose distribution volume was manually segmented and designated as ‘Control VOI’ at the ORN CECT. Subsequently, baseline, post-RT2, post-RT6 CECT scans were co-registered with ORN CECT using rigid registration algorithms of VelocityAI™ 3.0.1. Both ‘ORN’ & ‘Control’ VOIs were propagated from ORN CECT to other CECT scans at all three prior time points. (**Fig 2**)

**Fig 2.**
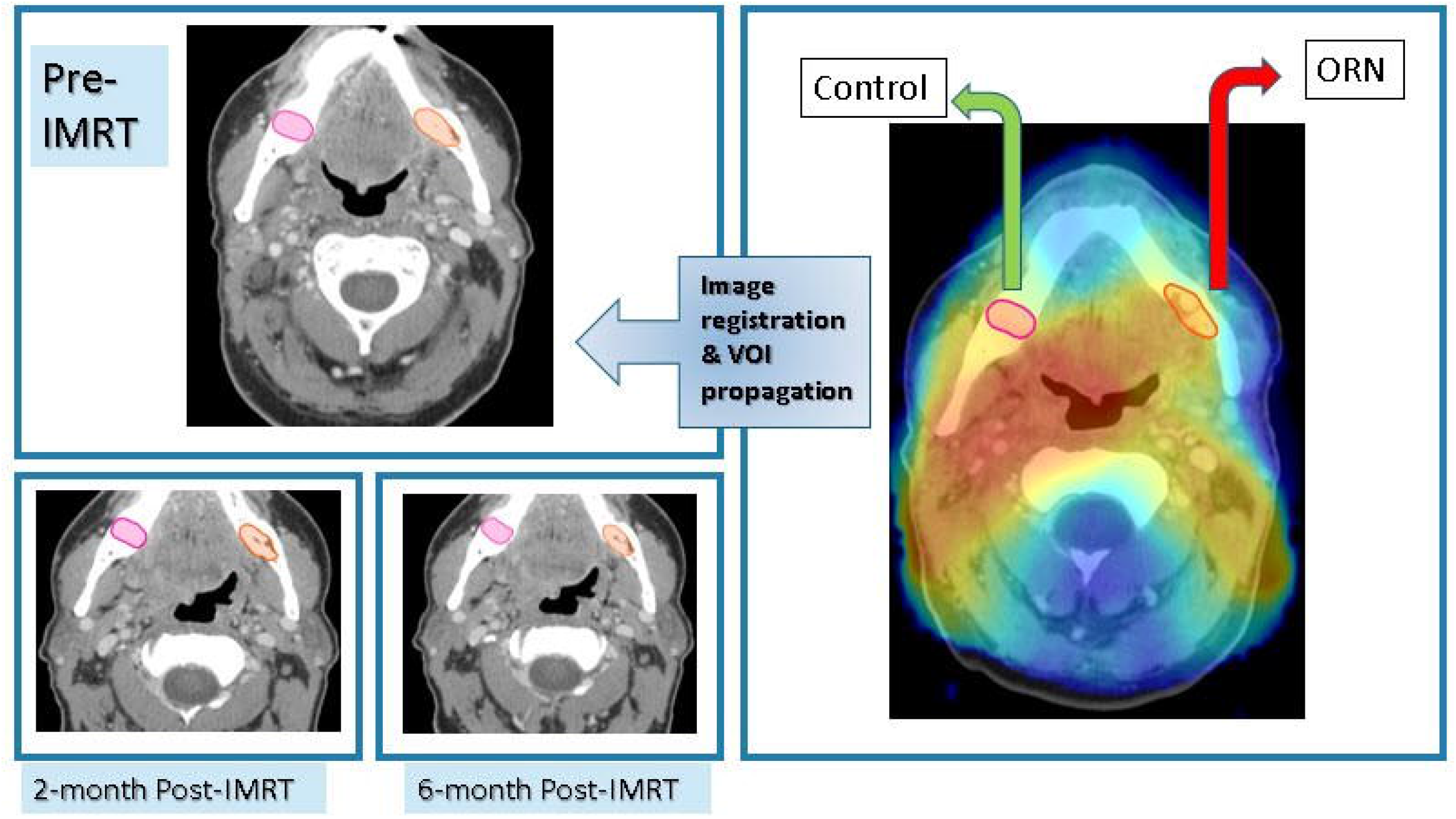
Imaging workflow. Registration of CECT scan at time of diagnosis of ORN to radiation dose grid as well as previous CECT scans at: baseline, 2-month and 6-month post-RT for each patient with subsequent propagation of ORN & ‘Control’ VOIs.

### Radiomics features extraction

Computed tomography scans with corresponding contoured VOIs were then extracted in the Digital Imaging and Communications in Medicine format (DICOM), as DICOM-RT and RT-STRUCT files, respectively. These files were then imported into an in-house image biomarker explorer (IBEX) software, built on MATLAB for subsequent radiomics feature extraction [15] along the same lines as previous studies [16, 17].

Radiomic features were derived from two VOIs that correspond to ORN and Control in the 3 prior time points: pre-IMRT, post-RT2 and post-RT6 CECT scans. The number of radiomic features extracted for each VOI summed up to 1645 individual features. They included a myriad of first- and second-order radiomics features, the latter calculated in both full 3-dimensional images (3D) as well as 2.5D, i.e. features calculated for each 2-dimensional slice and results were then combined. Other than shape features, a trilinear interpolation voxel resampling filter to 3 mm slice thickness and 1 mm^2^ pixel spacing was applied prior to feature extraction to standardize voxel size. First-order feature categories include: shape, intensity direct and intensity histogram. Whereas second-order feature categories encompass: Grey level co-occurrence matrix (GLCM), gray level run length matrix (GLRL) as well as neighborhood intensity difference. For GLCM and GLRL features, calculations from multiple spatial directions were combined to produce one value. For NID, 3 different permutations of neighborhood, i.e. 3, 5 or 7 were employed as in previous projects [18, 19].

### Radiomics features pre-selection and reduction

Initially, we worked with radiomic features computed from VOIs corresponding to ORN and Control for 24 patients. The number of radiomic features extracted for each patient is 1628. 3 patients did not have radiomic features computed for the post-RT6 time point and hence completely excluded from subsequent analysis. For these 21 patients, we only kept the radiomic features whose values are available for i) all 3 time points, and ii) both in ‘ORN’ and ‘Control’ VOIs. One patient has 2 distinct ORN lesions; accounting for a total number of 43 individual VOIs (22 ‘ORN’ and 21 ‘Control VOIs). Thus, we are then left with 1628 radiomic features from 43 VOIs, i.e. 22 ‘ORN’ and 21 ‘Control’.

Feature reduction by correlation was critical to ensure that the performance of any machine learning algorithm is not degraded because of very high degree of correlation in the features, or multicollinearity [20]. We first compute the Spearman correlation matrix [21] of the 1628 radiomic features at the pre-IMRT time point. We filter out the features whose average correlation level with all the remaining features is greater than a user-defined threshold. For our data, we used a threshold of 0.5. The threshold was chosen to reasonably balance the dual requirements of multicollinearity reduction and capturing data variation. Following correlation filtering, we reduced the number of features we analyze to 16 features. (**S1 Table**)

First –as a proof of concept-, we sought to establish that radiomics can quantitatively discriminate between ORN and non-ORN mandibular subvolumes. Mann-Whitney test [22] was used to identify specific radiomic features that show statistically significant differences between ORN and non-ORN high-risk VOIs.

### Functional Principal Component Analysis (FPCA)

Our hypothesis is that we can predict the risk of ORN by looking at the temporal evolution of radiomic features. A standard way of identifying temporal signatures in time series data is by using functional principal component analysis (FPCA) [23, 24]. FPCA takes multiple time series curves, as an input, and tries to find the underlying shape signatures that optimally can be used to represent all the curves. These shape signatures are called the functional Principal Components (PC). Each time series can now be represented by a weighted combination of each of the PCs. This technique has been used to predict outcomes from sequential data in a wide variety of fields such as remote sensing [25], stock markets [26], electroencephalogram (EEG) analysis [27], and cancer pathology [28]. Since our data is multivariate, in that we have a time series for multiple features for the same patient, we can compute the functional PCs for each feature. One way of representation would be to assume each feature is independent, concatenate the PC weights for each feature, and use this concatenated representation as input to a machine learning model. However, since each pair of features is correlated to various degrees, we use a technique called multivariate FPCA (MFPCA), which explicitly accounts for the relationship between the features [29-32]. We utilized the R package MFPCA for our temporal kinetics analysis [33].

The importance of FPCA is visually explained in **Fig 3**. We display 3 temporal trajectories from our data on the leftmost column. We observe that all 3 sequences *T*_1_, *T*_2_, and *T*_3_, have similar starting points. Further time series *T*_2_ and *T*_3_ have similar end points too. This mimics a significant scenario which we try to address, whereby neither the pre-radiotherapy features, nor the delta features can distinguish between the patients. However, FPCA is able to distinguish all 3, by accounting for both, the values taken by the time series, and the shape of the trajectory. The top 3 FPCs representing the dataset are shown visually in the top row. The relative contribution of each FPC to each of the time series is shown with arrows, the length of the arrows representing the magnitude, and green and red color indicating the sign (positive and negative, respectively) of the contributions. We can see that the magnitude and sign of the individual contributions from the PCs is quite different, and thus can help distinguish the three time series.

**Fig 3.**
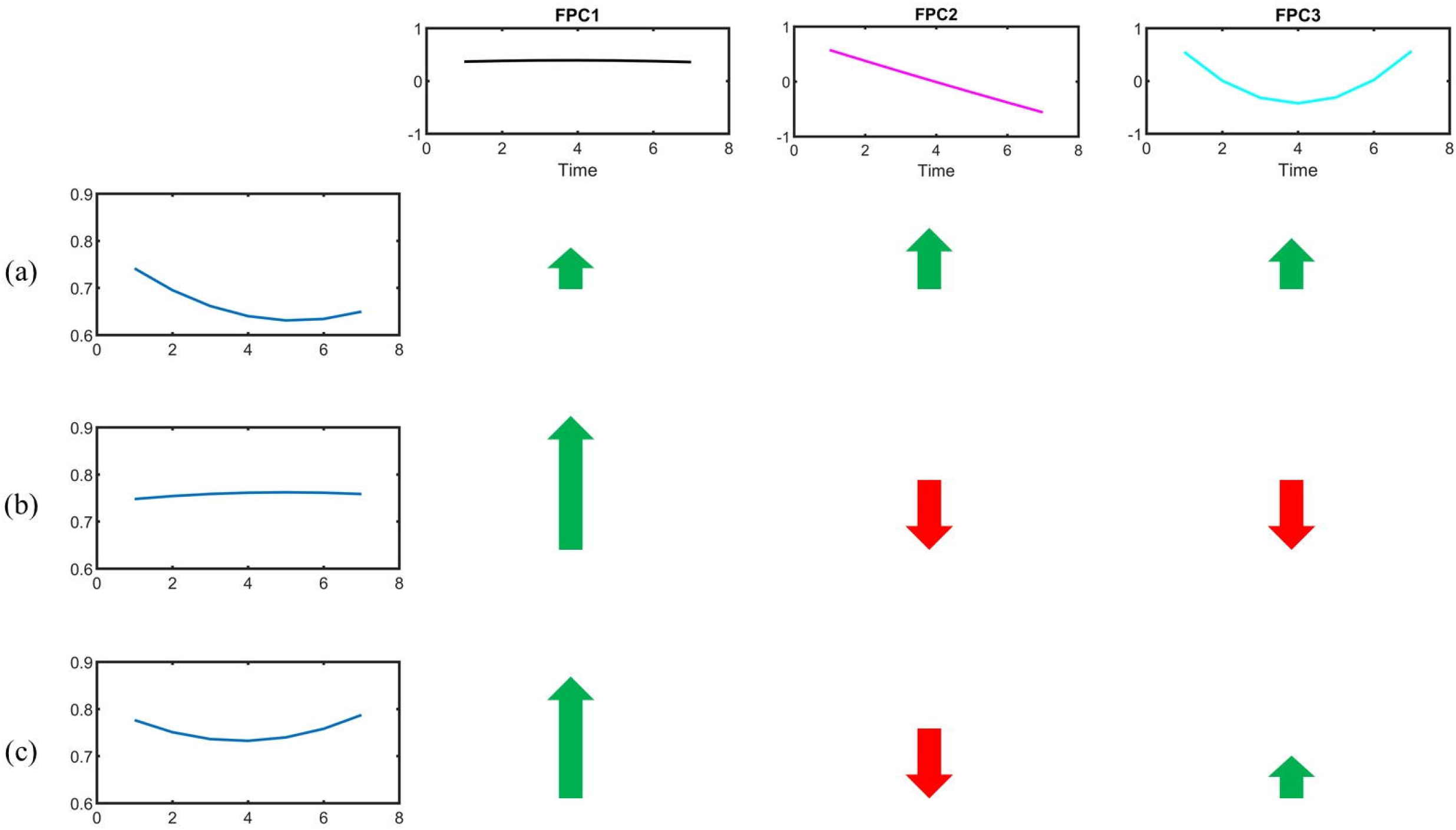
Visual explanation of the FPCA algorithm and its advantages. The first row displays the 3 functional principal components (FPCs). On the left column, the temporal evolution of a Gray Level Co-occurrence Matrix (GLCM)-3D feature is shown for three mandibular regions (a), (b) and (c). (a) and (b) did not develop ORN, while (c) did. We note that (a), (b) and (c) all have similar baseline values, so cannot be distinguished by a model built solely on pre-radiotherapy features. Further, (b) and (c) also have similar change in their values, which a delta radiomics model would see as equivalent scenarios. On the other hand, the difference in the temporal kinetics is efficiently encoded in the 3 FPCs. The color and length of the arrows indicate the sign (+ve or -ve) and magnitude (large or low) of relative contribution made by each FPC in explaining the time series. So, for example, regions (b) and (c), which appear alike to a pre-radiotherapy model and a delta radiomics model, can be readily distinguished because of the difference in relative contribution made by the 3^rd^ FPC.

### Training the random forest

Repeated random sampling to produce random forests [30] ensued where validation of each forest was performed using the left out observations, and the overall accuracy was calculated by averaging the class predictions of each of the forests. The random forest has been shown to be robust to overfitting and among the most effective of the commonly used classifiers. [34] Each forest used 500 trees, and each split was determined using 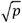 features where *p* is the number of features. The random forest calculations were performed using the random Forest package for R software [35]. To further examine the performance of the model, the ROC curves were plotted and the area under the curve (AUC) was calculated using pROC package for R [36].

## Results

### Patient information

Twenty-one patients with oropharyngeal cancer (OPC) were identified to have developed ORN subsequent to their definitive radiotherapy +/- chemotherapy course, either in induction and/or concurrent settings as in **Fig 1**. Eight patients developed grade 2 ORN, whereas 2 patients and 11 patients developed grade 3 and 4 ORN, respectively. Median time to ORN diagnosis was 20.3 months. **Table 1** represents patient demographics, tumor, radiation dose, and ORN disease characteristics.

**Table 1.** Patients, disease and treatment characteristics.

| **Characteristics** | **N (%)** |
| --- | --- |
| **Sex** |  |
| **Male** | 20 (95.2%) |
| **Female** | 1 (4.8%) |
| **Age at diagnosis, years: median (range)** | 61 (57-68) |
| **Ethnicity** |  |
| **White or Caucasian** | 17 (81%) |
| **Hispanic or Latino** | 2 (9.5%) |
| **African American** | 2 (9.5%) |
| **Smoking status** |  |
| **Current** | 10 (47.6%) |
| **Former** | 5 (23.8%) |
| **Never** | 6 (28.6%) |
| **Smoking pack-years (median; IQR)** | 10 (0-40.5) |
| **Tumor laterality** |  |
| **Right** | 9 (42.9%) |
| **Left** | 11 (52.4%) |
| **Midline** | 1 (4.7%) |
| **Oropharynx subsites** |  |
| **Base of tongue** | 12 (57.1%) |
| **Tonsil** | 7 (33.3%) |
| **NOS\*** | 2 (9.6%) |
| **T category** |  |
| **T1** | 2 (9.5) |
| **T2** | 10 (47.6%) |
| **T3** | 5 (23.8%) |
| **T4** | 4 (19.1%) |
| **N category** |  |
| **N0** | 2 (9.5%) |
| **N1** | 0 |
| **N2** | 19 (90.5%) |
| **N3** | 0 (0) |
| **Therapeutic combination** |  |
| **Induction chemotherapy (IC) followed by concurrent chemoradiation** | 10 (47.6%) |
| **IC followed by radiation alone** | 1 (4.8%) |
| **CC** | 10 (47.6%) |
| **Vital status** |  |
| **Alive** | 14 (66.7%) |
| **Dead** | 7 (33.3%) |
| **Radiation dose (median; IQR) [Gy]** | 70 (66-70) |
| **Radiation fractions (median; IQR)** | 33 (30-33) |
| **Onset of post-RT ORN (median; IQR)** | 20.3 (7.5-95) |
| **ORN laterality (in relation to primary tumor)** |  |
| **Ipsilateral** | 17 (81%) |
| **Contralateral** | 2 (9.5) |
| **Bilateral** | 2 (9.5%) |
| **Radiation dose at the ORN volume (median;** |  |
| **IQR) [Gy]** |  |
| **Mean dose** | 67.9 (59.5-71.2) |
| **Minimum dose** | 51 (44-59.4) |
| **Maximum dose** | 68.9 (67.6-73.1) |

IQR: inter-quartile range; Gy: Gray; NOS: Not otherwise specified; ORN: osteoradionecrosis

#### Radiomics can distinguish between ORN and non-ORN

An initial set of 1628 radiomic features were computed for each ORN and Control volume of interest (VOI) obtained from the 21 eligible patients across 3 time points of interest representing baseline (pre-IMRT), 2-month (post-RT2) post-IMRT, and 6-month (post-RT6) post-IMRT. Sixteen radiomics features were ultimately nominated as non-interrelated and consistently available for all three time points. As an initial exploratory step, we computed which of these 16 radiomic features were significantly different between the ORN and Control volumes of interest (VOIs) using a Mann-Whitney test. Furthermore, we also computed if each of these features is larger, or smaller, on average for the ORN VOI compared to the Control VOI. This demonstrates that certain radiomic features differ significantly between ORN and non-ORN regions, motivating us to investigate if their evolution can foretell ORN incidence. The significantly different features and their associated p-values are reported in **Table 2**.

**Table 2.**
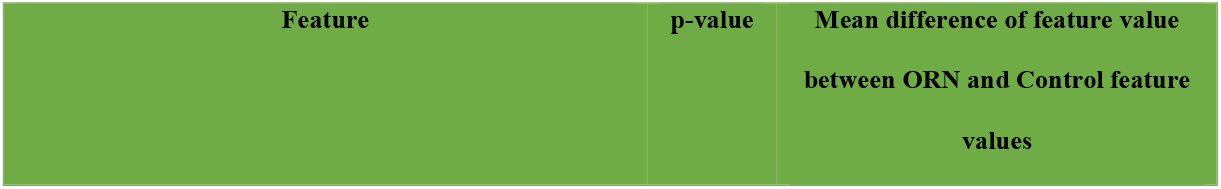
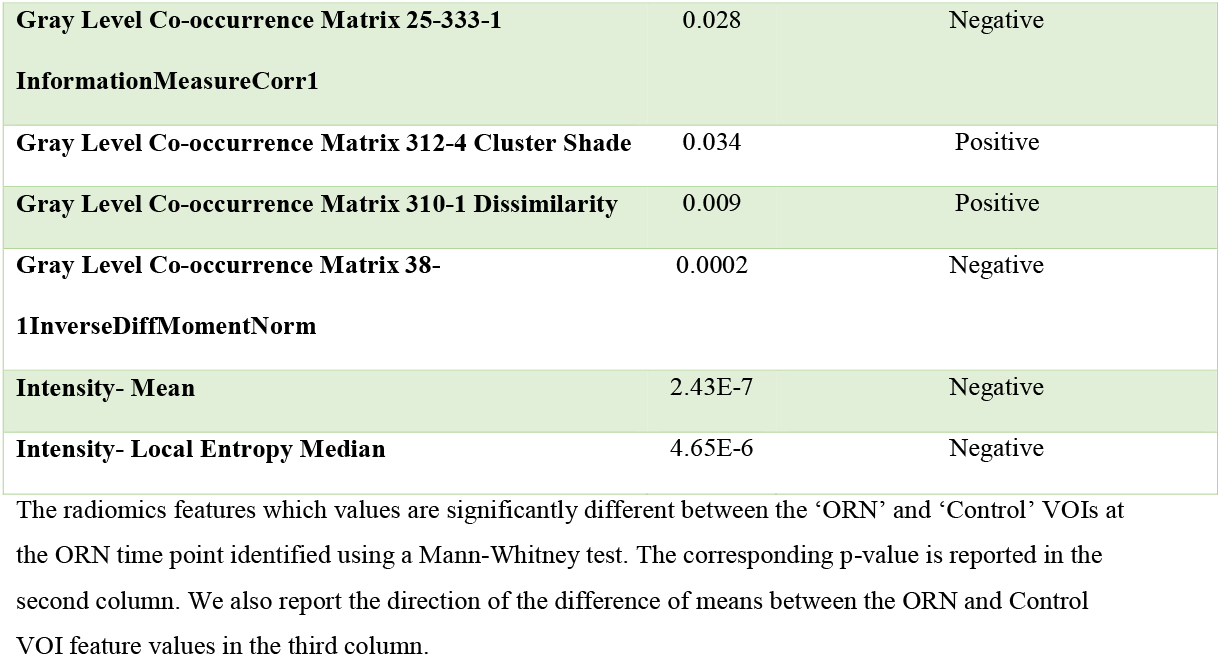
Significantly differing radiomics features between ORN and Control VOIs.

#### Model construction

We trained random forest models using 500 trees for each of multiple approaches as outlined below: (**Fig 4**)

**Fig 4.**
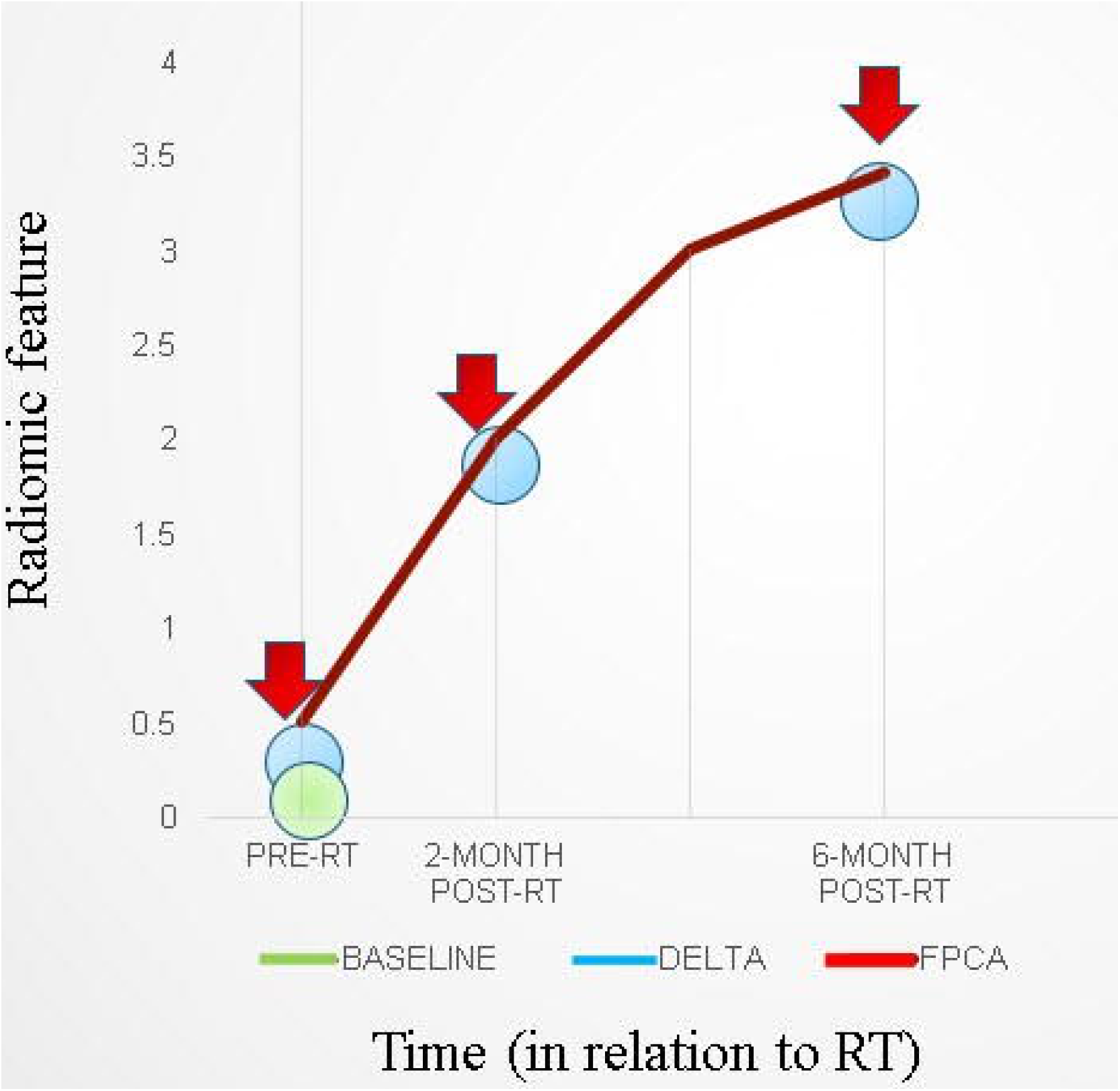
Overview of radiomics features based approaches. Various approaches to integrate radiomics features obtained at multiple (≥1) time points towards building predictive models.

- **Baseline:** Radiomic features computed on the pre-IMRT CECT scans.
- **Delta (2-month follow-up):** Relative change in the radiomic features from pre-IMRT to post-RT2
- **Delta (6-month follow-up):** Relative change in the radiomic features from pre-IMRT to post-RT6.
- **Temporal Trajectory:** The model built using the proposed multivariate functional principal component analysis (MFPCA) approach that models the temporal kinetics of the features. Since the time points are not uniformly spaced, we used cubic spline sequence completion to fill in radiomic features at intermediate monthly time points.
- **Baseline + Temporal Trajectory:** We combined the predictions from the baseline model and the temporal trajectory model to give a more robust ORN-risk predictor.

The corresponding areas under the curves (AUCs) and 95% confidence intervals (C.I.) for the prediction of occurrence of ORN ‘Yes vs No’, in both ‘ORN’ and ‘Control’ VOIs according to the 5 models are depicted in **Table 3** and illustrated in **Fig 5**. We noticed that the baseline features model gives an AUC of 0.59 (95% C.I: 0.41-0.76), while the temporal trajectory gives an AUC of 0.74 (95% C.I: 0.61-0.9). We further built an ensemble model that combines the predictions of the baseline model and the temporal trajectory model, to see if these two models have complementary information that improves performance. We achieved an AUC of 0.68 (95% C.I: 0.53-0.86), likely due to the poor performance of the baseline model which consequently was detrimental to the performance of the combined model. This suggests a more careful approach to choosing pre-IMRT features. Surprisingly, models constructed using percent changes ‘or delta changes’ of the radiomic feature values, performed poorly in predicting ORN incidence with AUCs of 0.64 (95% C.I: 0.46-0.81) and 0.56 (95% C.I: 0.39-0.74) for 2-month and 6-month delta changes, respectively. We further observe that the temporal trajectory and combined models have a consistent performance in both low-specificity and high-specificity regimes, in contrast to the delta models which performance is dependent on the regime of choice. This demonstrates that greater reliability is achieved by incorporating the temporal kinetics of the radiomic features.

**Table 3.**
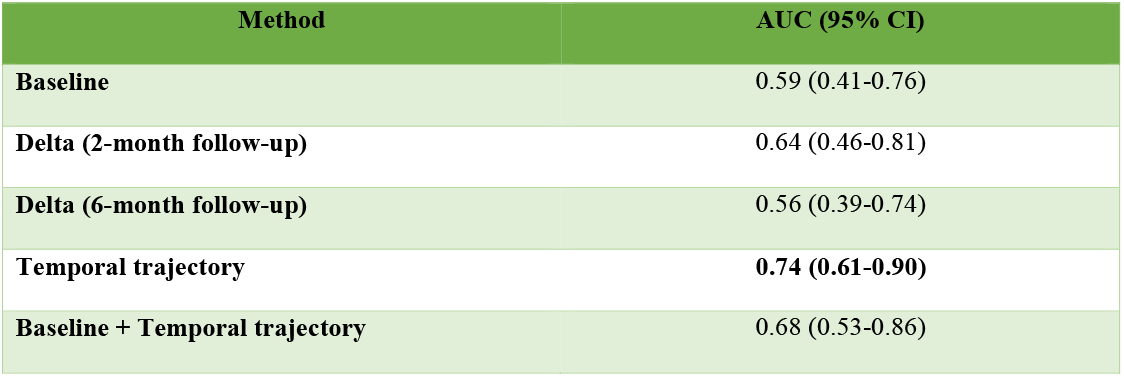
A comparison of the Areas under the curves (AUCs) and the 95% confidence intervals for the various approaches.

| Method | AUC (95% CI) |
| --- | --- |
| Baseline | 0.59 (0.41-0.76) |
| Delta (2-month follow-up) | 0.64 (0.46-0.81) |
| Delta (6-month follow-up) | 0.56 (0.39-0.74) |
| Temporal trajectory | **0.74 (0.61-0.90)** |
| Baseline + Temporal trajectory | 0.68 (0.53-0.86) |

**Fig 5.**
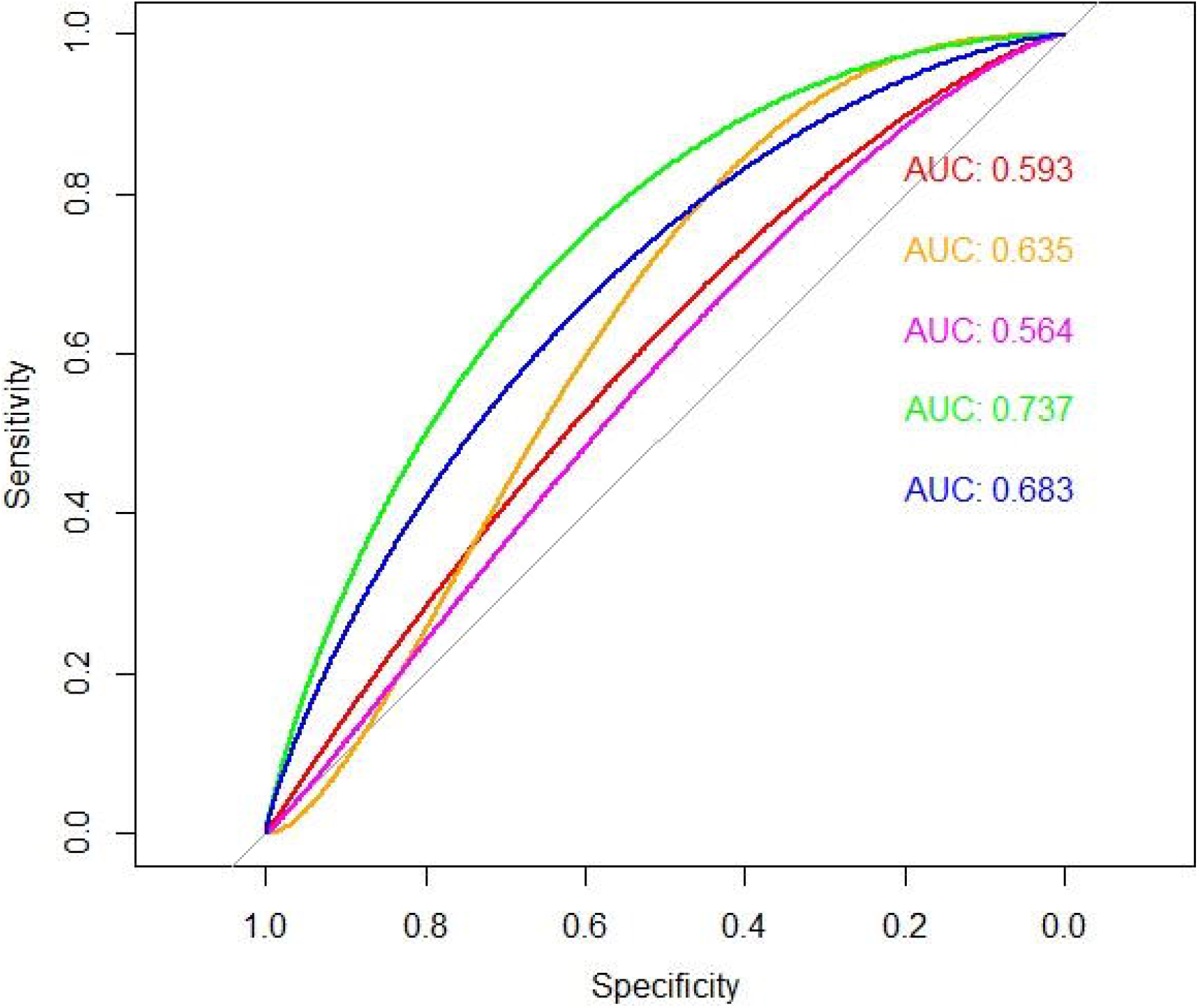
ROC curves computed for various radiomics feature based approaches. The temporal trajectory model using MFPCA (green) performs better than the other four models: i) baseline model (red), ii) delta model after 2-month follow-up (orange), iii) delta model after 6-month follow-up (purple), and iv) an ensemble of baseline and temporal trajectory models (blue).

## Discussion

The incidence of head and neck cancer is on the rise, despite reductions in smoking, owing to the recent prevalence of the human papillomavirus (HPV)-associated OPC epidemic [37]. Forward, it’s projected that hundreds of thousands of locally advanced OPC patients worldwide will receive radiation to the head and neck as a definitive treatment modality [38]. This rise in RT recipients implies that mandibular bone, which comprises the borders of the oropharynx, will be necessarily irradiated to ensure adequate tumor coverage with subsequently growing incidence of crippling sequelae such as ORN [39].

Osteoradionecrosis ranges from superficial, slowly progressive bone erosion/devitalization to pathological fracture in a previously irradiated field and may cause significant hardship in the afflicted individual [40, 41]. This is particularly apparent when considering devastating lifelong issues with oral hygiene, nutritional inadequacies, and difficulty with speech and resultant preclusion of social interaction [42]. Early diagnosis and intervention, whether conservative or surgical, are key for improving outcomes [43]. This essentially applies for grade II ORN, where no consensus has been reached regarding definitive treatment procedures [44, 45].

To date, no imaging modality/clinical nomogram have been shown to precisely foresee the potential risk of developing osteoradionecrosis following IMRT [2]. Being fully integrated throughout various phases of HNC management, sequential CECT scans via radiomics analytics can provide plethora of data that can serve as quantifiable surrogates of tissue vitality and vascularity, among others [46]. To our knowledge, this study is the first to characterize the kinetics of radiomics features of various mandibular subvolumes, before and after exposure to IMRT, to identify subvolumes at high risk ahead of developing ORN. Radiomics features were analyzed longitudinally for quantifying temporal changes in mandibular bone structure in a cohort of OPC patients.

This has been subsequently integrated into a framework for early prediction of ORN solely based on sequential diagnostic CECT scans. We implemented a Functional Principal Component Analysis (FPCA)- based approach that efficiently models the temporal evolution of radiomic features. The model built using a multivariate FPCA (MFPCA) representation of the entire temporal dataset, predicts the likelihood of ORN development with an AUC = 0.74 (95% C.I 0.61-0.9). We further built an ensemble model that combines the predictions of a baseline model built using pre-IMRT features, and the MFPCA-based model, in order to leverage information from both baseline feature values and temporal evolution of feature values, which achieved an AUC of 0.68 (0.53-0.86). This emulates the pathophysiology theories that combine pre-irradiation bone condition and RT-induced alterations on tissue, cellular and cytokine levels [47]. The latter involves: (1) endarteritis and vascular thrombosis with subsequent bone hypoxia and hypocellularity as well as atrophic fibrosis as a consequence of RT-induced activation and dysregulation of fibroblastic activity [44, 48]. The fact that the ensemble model does not perform better than the MFPCA-only model suggests the need to choose the pre-IMRT features in a way that is more clinically meaningful than a purely data-driven correlation thresholding approach.

Bone texture analysis has been investigated for years as a potential biomarker of a myriad of structural bone changes related to osteoporosis [49, 50]. Interestingly, **first-order** bone texture features derived from simulation CT scans were correlated to the risk of radiation-induced insufficiency fractures in patients undergoing pelvic radiation [51]. Along the same lines, for vascularization status, a previous study by Yin et al investigated the correlation between angiogenesis (or: new blood vessel formation) in primary renal cell carcinoma and radiomic imaging features from positron-emission tomography (PET) and/or MRI [52].

Our study identifies the bone radiomics features which temporal evolution is critical in determining ORN risk. These represent quantifiable imaging biomarkers that capture various intensity and spatial texture dimensions of the aforementioned RT-related bone environment changes in the irradiated field. Most of the discriminating features belong to: ‘Neighborhood intensity difference’ (NID) and ‘Grey level co-occurrence matrix’ (GLCM) categories. The GLCM is a matrix that expresses how combinations of discretized grey levels of neighboring pixels, or voxels in a 3D volume, are distributed along one of the image directions. Generally, the neighborhood for GLCM is a 26-connected neighborhood in 3D and an 8-connected neighborhood in 2D [53]. The ‘NID 2.5D Texture strength’ quantifies how uniform a texture is, i.e. complex textures are non-uniform and rapid changes in grey levels are common [54]. GLCM3 Cluster shade is a measure of the skewness or asymmetry of the matrix and is believed to be a more objective uniformity metric [55]. On the other hand, GLCM3 Contrast gauges grey level variations in the volume of interest, i.e. difference between the highest and the lowest values of a continuous set of pixels [56]. GLCM3 Correlation is a measure of texture smoothness, where higher values denote regions with similar gray-levels [57].

We have seen that there is significant information regarding ORN progression in the first 6 months after radiotherapy that can be robustly correlated to risk of ORN. Functional principal component analysis is an efficient statistical algorithm to capture the temporal evolution of the mandible landscape. Competing techniques such as pre-radiotherapy only models and delta radiomics models do not encapsulate how different features evolve with time. The FPCA efficiently encodes the temporal kinetics of the features into its functional principal components (FPCs). The radiomics data can now be compactly represented by only a small set of numbers, but can still capture its time-varying properties.

Furthermore, we implement a multivariate FPCA (MFPCA) that accounts for the correlations that exist between various radiomics features. MFPCA distils a large set of features to a few specific ones that encompass most of the data variation. This makes our prediction model more likely to generalize to new, unseen data [58]. We observe from the receiver operating characteristic curves that the temporal trajectory model performs consistently better than the other models in both the high- and low-specificity or false positive regions. This demonstrates the reliability of using temporal kinetics, for example, compared to a delta model, which we observed to have very different performance depending on the specificity value. The combined prediction model does not improve over the temporal trajectory only model, possibly because of the extremely poor performance of the baseline model. However, the combined model also performs consistently in both the low-false positive or high false positive regimes. We envisage that with a more careful choice of features, the baseline model can be improved, which will significantly improve the performance of the combined model.

The preliminary feature filtering step was performed by setting an upper limit of 0.5 on average correlation value for a given radiomic feature. Meaning, if a given radiomic feature correlated with all other features more than 0.5 on average, it was dropped from our feature set. The choice of value was made to whittle the number of features down from a mammoth 1628 to a more manageable 16. We also found that reducing the number of features further led to a drop in performance, which suggests loss of information crucial to prediction performance.

Our study accounted for the fact that artifacts from metal dental fillings are known to encumber target delineation and subsequent radiomics analysis [59, 60]. For this purpose, the presence of visible dental artifacts effect anywhere in the slices that encompassed ‘ORN’ or ‘Control’ VOIs at any time point precluded the integration of this scan and hence the patient’s data as an input to the model.

The fact that we excluded these patients with metal dental fillings, combined with the low event rate of ORN in the IMRT era, as well as the fact that we excluded patients with grade I ORN with no radiographically-evident bone lesions to delineate, contributed to the low sample size; hence limiting the generalizability of the resulting model. The small sample size limited us to apply automatically generated radiomics features instead of engineering features that are explicit surrogates for early vascular injuries of the mandible. Another limitation of this study is the conceivable uncertainties introduced from varied acquisition parameters or incongruence among various scanners, or even between different models from the same vendor [61]. Most patients had their scan performed at our center along the same acquisition parameters. Moreover, we have applied a pre-processing trilinear interpolation aiming at standardizing voxel size to reduce or eliminate relevant variability in radiomics features [62]. The results also suggested that the performance changed rapidly when we changed the number of features, which suggests the need for a more careful feature-filtering algorithm. Designating a ‘Control’ VOI that share the same image, time point, and deposited radiation dose with the ‘ORN’ VOI is an approach we have used and would recommend for future multi-institutional radiomics studies.

## Future Directions

Not far from longitudinal imaging studies, our team previously showed that Dynamic Contrast-Enhanced (DCE-MRI) can provide biomarkers that are physiological correlates of acute mandibular vascular injury and recovery temporal kinetics [63]. This has further motivated a National Institute of Dental and Craniofacial Research (NIDCR)-funded prospective trial that explores correlation between DCE-MRI derived spatiotemporal parameter maps following external beam radiation therapy (EBRT) and subsequent development of ORN [64]. Our results may prompt the investigation of DCE-MRI-derived radiomics analytics and subsequent integration into the overall predictive model; thus, providing more data inputs for the machine learning techniques tested.

Furthermore, the availability of larger cohorts will provide potential avenues for model validation and generalization over the whole mandible in patients with ORN versus healthy controls. A proposed application would be engineering radiomics features that are explicit surrogates for osteoclastic dysregulation and subsequent fibro-atrophic bone changes, and maybe monitoring the response to common therapeutic maneuvers, such as pentoxyfilline.

## Conclusion

Radiomics analysis allows for quantification of changes in RT-related bone structure from diagnostic imaging modalities with subsequent integration of serially derived radiomics features into an ORN probability computational tool. Computationally, FPCA efficiently encodes the temporal kinetics of a given radiomic feature. The MFPCA then compactly combines the temporal information from FPCA from multiple radiomic features.

In summary, we hope this study calls professionals’ attention to non-traditional inputs (radiomics), dimensions (temporal kinetics), and innovative statistical approaches (MFPCA) to improve interpretation and integration of imaging biomarkers into RT toxicities prediction and mitigation. In this work, we have thus provided an end-to-end framework for predicting the risk of RT-related ORN based entirely on radiomic features.

## Data Availability

Clinical dataset is not available as it includes personal health identifiers (PHI). It is possible for de-identified data to be made available upon reasonable request. 

## Supporting information

**S1 Table. Filtered radiomic feature set**. The final set of 16 features chosen after correlation filtering, which are used for building baseline, delta, and FPCA models.

